# Multilevel associations between skills use, engagement, and treatment outcome in self-guided internet-delivered dialectical behavior therapy for substance use disorders

**DOI:** 10.1101/2025.10.07.25337533

**Authors:** Danielle Downie, Alexander R. Daros, Chelsey R. Wilks, Lena C. Quilty

**Affiliations:** Department of Psychological Clinical Science, University of Toronto, Toronto, Ontario, Canada; Campbell Family Mental Health Research Institute, Centre for Addiction and Mental Health, Toronto, Ontario, Canada; Department of Psychology, University of Windsor, Windsor, Ontario, Canada; Tori Health, Los Angeles, CA, USA; Department of Psychiatry, University of Toronto, Ontario, Canada

**Keywords:** dialectical behavior therapy, substance use disorders, multilevel modeling, emotion dysregulation, mindfulness, engagement

## Abstract

Internet-delivered dialectical behavior therapy (iDBT) represents a scalable and accessible treatment approach for individuals with substance use disorders (SUDs), yet little is known about the mechanisms of change and the role of engagement in this format. This study examined within- and between-person associations between changes in skills-based constructs (mindfulness, DBT skills use, emotion dysregulation) and two treatment outcomes (SUD severity and functional disability) across a 12-week self-guided iDBT program. The moderating role of treatment engagement was also evaluated. 72 participants with past year SUDs were randomized to immediate or delayed iDBT and completed assessments at baseline and weeks 4, 8, and 12. Multilevel models were used to assess within- and between-person associations between skills and outcomes over time. Several indices of engagement were used including treatment acceptability, usability, and hours/days spent on iDBT. SUD severity and functional disability significantly decreased over the 12-week period. Within-person increases in emotion dysregulation were associated with higher SUD severity and disability, while higher levels of mindfulness between-persons were associated with lower disability. DBT skills were not significantly associated with outcomes. Greater within-person treatment acceptability (cognitive engagement) moderated reductions in SUD severity over time. Findings support the role of emotion dysregulation and mindfulness emerging as key correlates of treatment response. Treatment perceptions such as acceptability and usefulness may enhance outcomes, along with behavioural engagement. Future work should refine measurement of DBT skill acquisition and investigate longer-term functional impacts of digital DBT interventions.

**Author Summary:** In this study, we sought to understand potential mechanisms mechanisms of change in self-guided internet-delivered dialectical behavioural therapy (DBT) for individuals with substance use disorders. Using multilevel modeling, we evaluated the within- and between-person variation in three targets (mindfulness, DBT skills, and emotion dysregulation) over the treatment and follow-up period of 12 weeks and examined their association with treatment response (defined as substance use severity and functional disability). We then examined cognitive and behavioral facets of engagement as moderators of treatment response as well. With few DBT digital tools in existance, this study provides preliminary evidence that self-guided internet-delivered DBT tools can improve treatment targets similar to standard individual and group DBT formats. Overall, the study provides novel evidence on how several variables may enhance treatment outcome for individuals with substance use disorders, thereby informing future digital intervention trials.

## Introduction

Substance use disorders (SUDs), which involve the problematic use of alcohol, nicotine, cannabis, and other substances, are a pervasive global health concern with severe impacts on individuals, families, and society at large [1]. SUDs are associated with a high degree of functional disability, including impaired relationships, employment, and overall functioning [2]. Broad and scalable intervention efforts are critical for reducing long-term harms. International institutions emphasize the need for equitable access to treatment and harm reduction services to combat this ongoing public health issue [3,4]. However, research suggests that fewer than 10% of people with SUDs receive specialized treatment, and of those who receive care, dropout rates often exceed 30% for psychological interventions [5]. These challenges highlight the necessity of improving access to treatment as well as engagement with them, to ensure interventions meet the diverse needs of individuals with SUD.

There is a long-standing recognition that substance use functions to regulate emotions [6]. This regulatory function leads to reinforcement and maintenance of substance use [7]. Given the prominent role of emotion regulation in mental disorders often co-occur with harmful substance use, such as mood and anxiety disorders [8], psychological interventions that reduce harmful substance use by developing a range of healthy emotion regulation skills represent promising transdiagnostic treatment approaches that could improve not only SUDs but also conditions that often co-occur with SUDs. Dialectical behavior therapy (DBT) was initially developed for the treatment of individuals with severe suicide risk and individuals with borderline personality disorder (BPD) [9,10]. Due to the high number of individuals with BPD presenting with co-occurring SUDs, specialized DBT content was created to target and address harmful substance use [11]. DBT for individuals with SUDs addresses intense emotions (e.g., craving, withdrawal), impulsive behaviors (e.g., managing urges, avoiding cues), and interpersonal effectiveness (e.g., increasing community involvement) using a skills-based approach. The goal of DBT for SUDs is to reduce harmful substance use and dysfunctional behaviours while increasing behavioural control [11,12].

Research supports the feasibility and efficacy of DBT for SUDs, wherein DBT includes the fulsome offering of in-person group and individual therapy sessions, along with phone coaching in crisis situations and consultation for therapists. This comprehensive DBT protocol is associated with decreased frequency substance use, improved social functioning, and enhanced emotion regulation in early trials in those with SUDs with or without co-occurring BPD [13–17]. Additional research has supported the use of stand-alone group-delivered DBT skills training (often abbreviated as DBT-ST) for SUDs [18–20]. One recent systematic review of DBT-ST for SUDs reported reduced substance use and improved emotion regulation within participants over treatment [21]. Collectively, results support both comprehensive DBT and skills training formats to achieve positive outcomes for individuals with SUDs. To further increase the reach of DBT, researchers have developed mobile and internet-delivered formats to study its impact in SUDs using a scalable format that increases accessibility. One recent systematic review of DBT-ST for SUDs reported reduced substance use and improved emotion regulation within participants over treatment [21]. Collectively, results support both comprehensive DBT and skills training formats to achieve positive outcomes for individuals with SUDs. To further increase the reach of DBT, researchers have developed mobile and internet-delivered formats to study its impact in SUDs using a scalable format that increases accessibility.

Mobile and internet-delivered DBT (often abbreviated as iDBT) offers an increasingly accessible and flexible approach to care. This delivery method has gained traction over the past decade due to high demands for mental health treatment (compounded by the COVID-19 pandemic) and the challenges in attending in-person treatments such as comprehensive DBT [22,23]. The first randomized controlled trial of self-guided iDBT for harmful substance use recruited individuals with high suicidality and heavy alcohol use [24]. Individuals were randomized to immediate or delayed access, with roughly 85% of participants completing the first module and one-third completing all eight. Those who remained in the study longer and completed more modules reported significantly reduced suicidal ideation, alcohol quantity and severity, and emotion dysregulation over 16 weeks. Schroeder and colleagues [25] demonstrated feasibility and acceptability for the first iDBT mobile app, called *Pocket Skills,* as an adjunct to standard DBT treatment in a non-randomized trial of participants with mostly depressive and anxiety disorders. The sample consisted of 73 individuals with participants meeting criteria for depression (n=38), generalized anxiety disorder (n=35), BPD (n=29), post-traumatic stress disorder (n=20), bipolar disorder (n=10), and other disorders. After 4 weeks in the study, the authors reported decreased depression and anxiety symptoms, while also increasing DBT skills use. Results suggested that *Pocket Skills* helped participants engage in their DBT and practice and implement skills in their environmental context due to its mobile nature.

*Pocket Skills* was revised and investigated as a self-guided intervention for SUDs in a subsequent randomized controlled trial [26]. Specifically, individuals with SUDs were randomized to receive access to the intervention immediately or after a 4-week delay and were followed for 12 weeks total. In addition to being feasible and acceptable, participants reported reductions in SUD severity, depression, anxiety, suicidality, functional disability, and emotion dysregulation in both the immediate and delayed groups. DBT skills as well as mindful awareness improved in both groups, with a higher degree of engagement reported within the immediate versus the delayed iDBT group. Overall, results support the potential of iDBT in treating SUDs in a complex sample with a high degree of concurrent mental health disorders.

Research on the potential mechanisms underlying DBT outcomes have typically focused on gains in mindfulness, emotion regulation ability, distress tolerance, and DBT skills [27]. Increases in DBT skills use were associated with improved treatment outcomes in a transdiagnostic DBT group treatment, suggesting that skills use may be a mediator of treatment outcomes in DBT more broadly [28,29]. In a sample of adolescents undergoing DBT, both improvements in emotion regulation and interpersonal sensitivity were significant predictors of change in depression and anxiety [30]. Participants in this sample included youth aged 14 to 25 who had a history of suicide attempts in the last 5 years. Additional research support improvements in emotion regulation and DBT skills as mediator of suicidality and self-harm reductions in youth undergoing DBT for suicidal behaviors and self-harm [31]. Given that individuals will vary in their mindfulness, emotion regulation, and DBT skills at the outset of an intervention, and have different improvement trajectories, it is important to consider the between- and within-person variation of these variables over the course of treatment.

Engagement in treatment is also critical to clinical outcomes and yet, it remains an ill-defined construct. In traditional psychotherapy, engagement can be measured as attendance and participation in treatment, as well as homework completion, and is often associated with improved outcomes [32,33]. Within a standard DBT context, increased weekly homework completion and participation in phone coaching predicted improved suicidal behaviors and substance use in an outpatient program [34]. With the proliferation of digital health interventions, there have been attempts to conceptualize and define engagement given its association with treatment outcome [35,36]. Rather than focusing only on behavioral engagement metrics (e.g., frequency of use and time spent on the intervention), Kelder and colleagues [35] propose cognitive components as well, such as perceptions about the intervention (e.g., its acceptability and usability). In one study of patients with SUDs, the number of modules completed within an online self-guided cognitive behavioral therapy treatment program (e.g., behavioral engagement), was positively associated with reduced substance use [37]. Within the limited iDBT literature, only one study examined the relationship between behavioural engagement and treatment outcomes. However, against expectations, neither module completion nor diary card entries within an iDBT app were predictive of disordered eating symptom improvement [38]. This literature gap provides the opportunity to further understand the relationship between online iDBT treatment engagement and treatment outcome in SUDs.

### The Present Study

The current study sought to extend the results of a previous trial by examining predictors of treatment outcomes (i.e., SUD severity and functional disability) in individuals with SUDs who received iDBT. Considering the literature on both traditional and internet-delivered DBT formats, we examined whether changes in DBT skills, mindfulness, and emotion dysregulation predicted response to treatment for individuals with SUDs. Using multilevel modelling, we considered both the within- and between-person associations between skills-based variables and treatment outcomes, while also considering key demographic and control covariates. We then further examined the moderation of iDBT treatment outcomes by examining several indices of cognitive and behavioural engagement: treatment acceptability, usability (defined as ease of use, interface satisfaction, and usefulness), and time spent on the intervention (in hours and unique days accessed). We hypothesized that (1a) within-person increases in mindfulness and DBT skills would be negatively associated with SUD severity and functional disability over time, while (1b) within-person decreases emotion dysregulation would be positively associated. We further hypothesized that (2a) higher between-person mindfulness and DBT skills would be negatively associated with SUD severity and functional disability, whereas (2b) higher emotion dysregulation would be positively associated. Finally, we hypothesized (3a) that we would find evidence for moderation for time-varying, cognitive engagement variables (e.g., treatment acceptability and usability) at the within-person level and (3b) behavioural engagement (e.g., hours/days spent on iDBT) at the between-person level, such that higher engagement would strengthen the relationships between time and treatment outcome.

## Methods

### Study Design and Recruitment

This study is a secondary analysis of a preregistered, two-arm, single-blinded, parallel-group randomized controlled trial of iDBT for SUDs (NCT05094440; [26]). The research questions and analyses presented in the current study are unique and do not overlap with previous work. Participants were divided into two groups: those receiving immediate access to iDBT and those assigned to a waitlist for four weeks before receiving access. Participant recruitment occurred from August 2022 to March 2023, and utilized referrals from psychiatric hospital clinicians, waitlists, and research registries, as well as self-referrals from local community advertisements on hospital websites, social media, private clinics, and community organizations. Advertisements targeted individuals seeking to reduce alcohol or substance use, through an internet-delivered intervention provided at no cost. Prospective participants were informed about the study and underwent phone-based pre-screening for eligibility. To be included in the study, participants had to be between the ages of 18–65 years; fluent in English; willing to complete study requirements; not currently enrolled in CBT or DBT (peer support and psychiatric services were permitted); had a past year SUD; had internet access and literacy; and at least a score of 4 on a modified Contemplation Ladder question (indicating moderate readiness to reduce substance use). Exclusion criteria included practical barriers to participation (e.g., extended absences); acute psychiatric conditions (e.g., suicidality, psychotic disorder) or medical conditions requiring immediate attention; and enrollment in other psychological intervention studies. Use of psychotropic medications did not disqualify participation.

### Procedure

Eligible participants attended a 45-minute baseline session via videoconference to provide electronic informed consent, complete a demographic survey, and participate in a semi-structured diagnostic interview. Participants were then randomized into immediate or delayed iDBT groups. Participants in the immediate group received the iDBT website URL and an access code, completing the sign-in process during the session. The delayed access group completed the sign-in process after four weeks in a second, brief videoconference session. Both groups were instructed to spend at least 1-2 hours per week over the first 4 weeks, which was defined as the acute treatment period. Engagement was captured up to 12 weeks post-baseline for both groups.

Questionnaire assessments were conducted at baseline and then 4-, 8-, and 12-weeks post-baseline, using REDCap (Research Electronic Data Capture) [39] and distributed via email or text every four weeks. Automated reminders were sent daily for up to four days, beginning two days before the due date. Additional calls or meetings with the experimenter were offered on an as needed basis for technological support. Participants were compensated an average CAD $59 (maximum CAD $70) for baseline and follow-up assessment completion.

### iDBT Intervention

*Pocket Skills 2.0* is an iDBT intervention developed by author CRW in collaboration with Microsoft Research and Marsha Linehan, the creator of DBT. It was built upon the most recent DBT manual available [40] and uses a web-based portal built on the Microsoft Azure platform that is compatible with any internet browser in addition to the Android and iOS mobile operating systems. This iDBT intervention incorporates lessons following the core modules of DBT (mindfulness, emotion regulation, distress tolerance, and interpersonal effectiveness) as well as a specific module focused on addiction. Within each module, participants select specific skills and are presented with a brief video featuring Dr. Linehan introducing the skill and its uses. A practice session then ensues with the rule-based chatbot, which allows for feedback through both open-ended text input and a closed selection of responses. The chatbot guides users on how to select skills to use in different situations that may arise as well as the ability to gain points and unlock additional content, which increases user engagement.

### Measures

Demographic information was collected during a brief clinical interview at the baseline meeting. The Diagnostic Assessment and Research Tool (DART; [41]) is a brief semi-structured diagnostic interview that was used to assess depressive, anxiety, bipolar, obsessive-compulsive, trauma and stressor, psychotic, and SUDs according to the *Diagnostic and Statistical Manual of Mental Disorders, Fifth edition* [42]. All interviews were completed by the second author, who is a licensed clinical psychologist. Rates of current mental disorders were as follows: Major Depressive Disorder (51.4%), Persistent Depressive Disorder (25.0%), Bipolar I/II Disorder (8.3%), Panic Disorder (5.6%), Agoraphobia (8.3%), Specific Phobia (4.2%), Generalized Anxiety Disorder (51.4%), Social Anxiety Disorder (30.6%), Post-Traumatic Stress Disorder (25.0%), Alcohol Use Disorder (65.3%), Cannabis Use Disorder (33.3%), Nicotine Use Disorder (20.8%), and Stimulant Use Disorder (12.5%).

### Skills-Based and Outcome Measures

Reliability coefficients for these measures are listed in Table 1.

**Table 1.**
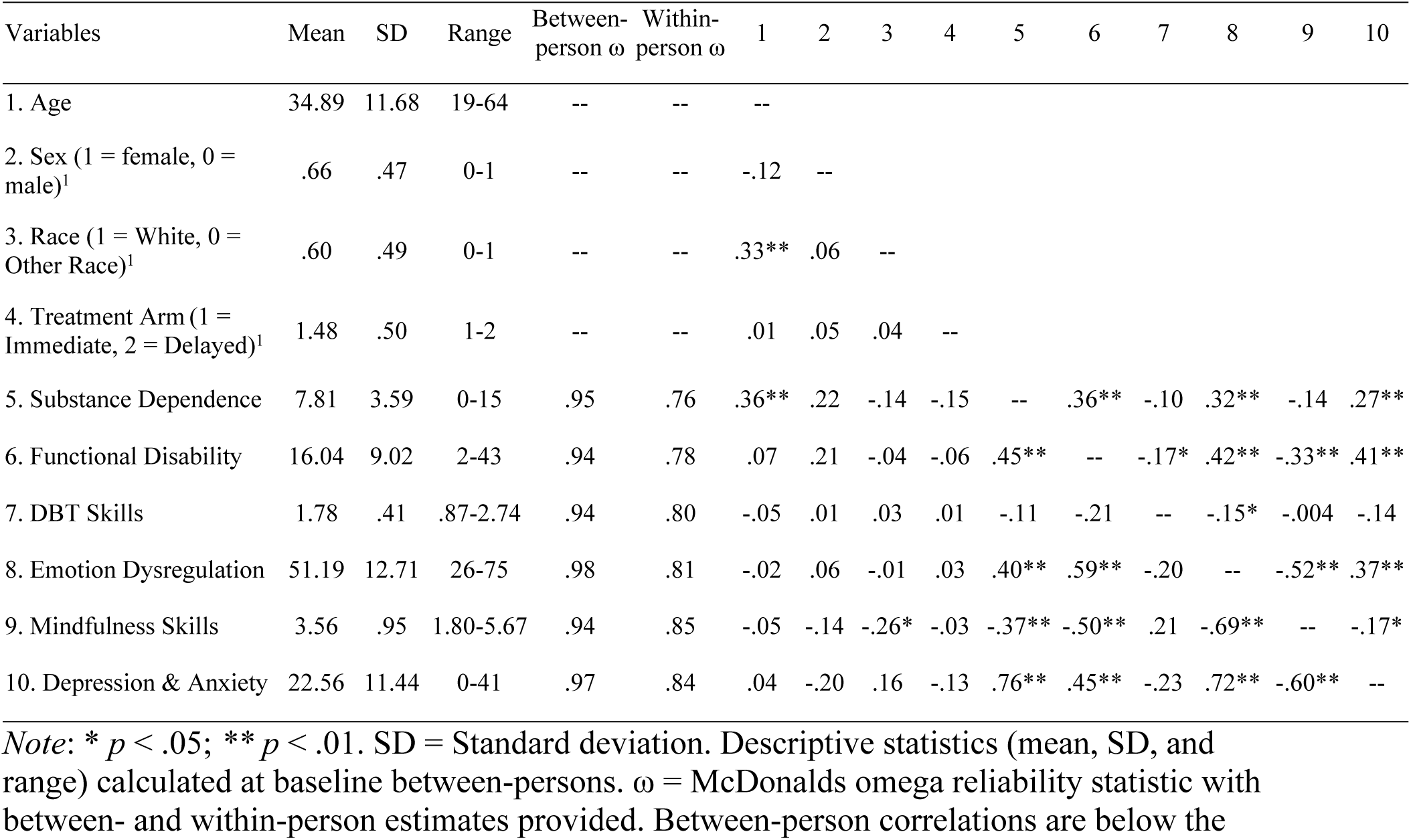
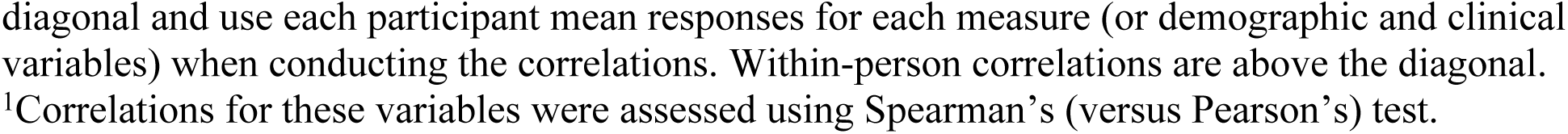
Baseline Descriptive Statistics and Between- and Within-Persons Correlations for Demographic and Variables of Interest.

| Variables | Mean | SD | Range | Between-person $\omega$ | Within-person $\omega$ | 1 | 2 | 3 | 4 | 5 | 6 | 7 | 8 | 9 | 10 |
| --- | --- | --- | --- | --- | --- | --- | --- | --- | --- | --- | --- | --- | --- | --- | --- |
| 1. Age | 34.89 | 11.68 | 19-64 | -- | -- | -- |  |  |  |  |  |  |  |  |  |
| 2. Sex (1 = female, 0 = male)<sup>1</sup> | .66 | .47 | 0-1 | -- | -- | -.12 | -- |  |  |  |  |  |  |  |  |
| 3. Race (1 = White, 0 = Other Race)<sup>1</sup> | .60 | .49 | 0-1 | -- | -- | .33** | .06 | -- |  |  |  |  |  |  |  |
| 4. Treatment Arm (1 = Immediate, 2 = Delayed)<sup>1</sup> | 1.48 | .50 | 1-2 | -- | -- | .01 | .05 | .04 | -- |  |  |  |  |  |  |
| 5. Substance Dependence | 7.81 | 3.59 | 0-15 | .95 | .76 | .36** | .22 | -.14 | -.15 | -- | .36** | -.10 | .32** | -.14 | .27** |
| 6. Functional Disability | 16.04 | 9.02 | 2-43 | .94 | .78 | .07 | .21 | -.04 | -.06 | .45** | -- | -.17* | .42** | -.33** | .41** |
| 7. DBT Skills | 1.78 | .41 | .87-2.74 | .94 | .80 | -.05 | .01 | .03 | .01 | -.11 | -.21 | -- | -.15* | -.004 | -.14 |
| 8. Emotion Dysregulation | 51.19 | 12.71 | 26-75 | .98 | .81 | -.02 | .06 | -.01 | .03 | .40** | .59** | -.20 | -- | -.52** | .37** |
| 9. Mindfulness Skills | 3.56 | .95 | 1.80-5.67 | .94 | .85 | -.05 | -.14 | -.26* | -.03 | -.37** | -.50** | .21 | -.69** | -- | -.17* |
| 10. Depression & Anxiety | 22.56 | 11.44 | 0-41 | .97 | .84 | .04 | -.20 | .16 | -.13 | .76** | .45** | -.23 | .72** | -.60** | -- |

Note: \* $p < .05$ ; \*\* $p < .01$ . SD = Standard deviation. Descriptive statistics (mean, SD, and range) calculated at baseline between-persons. $\omega$ = McDonalds omega reliability statistic with between- and within-person estimates provided. Between-person correlations are below the
diagonal and use each participant mean responses for each measure (or demographic and clinical variables) when conducting the correlations. Within-person correlations are above the diagonal. <sup>1</sup>Correlations for these variables were assessed using Spearman's (versus Pearson's) test.

The *Mindful Attention and Awareness Scale* [43] is a 15-item measure of mindful attention and awareness, which is a primary target of DBT skills training. Items were rated on a 6-point Likert scale (1=Almost Never to 6=Almost Always). Higher scores reflect higher levels of dispositional mindfulness ability.

The *Difficulties in Emotion Regulation Scale* [44] is an 18-item self-report measure of difficulties in regulating emotions, including six subscales: emotional awareness, emotional clarity, difficulties in goal orientation, difficulties in impulsivity, nonacceptance, and difficulties selecting strategies. Items are rated on a 5-point Likert scale (1=Almost Never to 5=Almost Always). Higher scores indicate greater problems in emotion regulation, and lower scores are indicative of greater skills and ability in managing emotions.

The *DBT Ways of Coping Checklist* [45] is a 59-item self-report measure that assesses the frequency of maladaptive and adaptive skills used to manage difficult situations over the past month, with good internal consistency and test-retest reliability. In this study, we used the 38-item adaptive skills subscale, which includes skillful behaviors often learned in DBT without using DBT-specific language. Items were rated on a 4-point Likert scale (0=Never Used to 3=Regularly Used) and higher scores reflect more adaptive skills usage.

The *Severity of Dependence Scale* [46] is a 5-item measure of SUD severity that is well-validated against substance use disorders. Items are rated on a 4-point Likert scale (0=Never/Almost never to 3=Always/Nearly always), with higher scores indicating a higher level of substance dependence symptoms. Participants were first asked to indicate which class of substance (including alcohol) they were experiencing the most difficulties with.

The *World Health Organization Disability Assessment Schedule* (Short Form 2.0; [47]) is a 12-item measure assessing six domains related to quality of life: physical health, psychological, social relationships and environment. Items were rated on a 5-point Likert scale (0=None to 4=Extreme or cannot do) with higher scores indicating greater functional disability.

The *Patient Health Questionnaire, Depression Subscale* [48] is a 9-item measure used to baseline depressive symptoms over the past two weeks. The *Generalized Anxiety Disorder-7 Scale* [49] is a 7-item measure used to assess generalized anxiety symptoms over the past two weeks. For both measures, items are rated on a 4-point Likert scale (0=Not at all to 3=Nearly every day). To create a sensitivity analysis in contrast to SUD severity and functional disability, while also avoiding multicollinearity (baseline *r* = .77), we summed the respective scores to create one composite variable, a previously validated procedure [50].

### Engagement Measures

The *Treatment Acceptability Questionnaire* [51] is a 6-item self-report scale that was administered during follow-up assessments to examine perceived acceptability, safety, and trustworthiness using a 7-point Likert scale from 1 to 7 (anchors differ per item). A total score is created between 7 and 42, with higher scores indicating greater acceptability. In the current study, the between/within ω reliability was .92/.55.

The *mHealth App Usability Questionnaire* [52] is an 18-item scale that was administered during follow-up assessments to examine perceived ease of use (between/within ω reliability = .91/.75), interface and satisfaction (between/within ω reliability = .86/.73), and usefulness (between/within ω reliability = .89/.72) of the iDBT platform. Items are rated on a 7-point Likert scale from 1 (strongly agree) to 7 (strongly disagree), with lower scores suggesting better ratings.

Other measures of engagement included the total amount of time spent on the application (in hours) as well as the number of unique days where participants recorded at least 5 minutes of activity. These measures were calculated based on the duration of the trial (i.e., 12 weeks).

### Statistical Analyses

We first collapsed data across treatment arms given that there were no significant group differences in our treatment outcomes of interest (see [26]). Instead, treatment arm was added as a covariate in all analyses. Data were organized as multiple observations nested within individual participants. A total of 261 surveys were attempted across all participants and weeks, which consisted of both baseline (time 0) and follow-up surveys at weeks 4, 8, and 12. No data was missing from the baseline, and participants returned at least partially completed follow-up questionnaires at rates of 94% (week 4), 78% (week 8), and 81% (week 12). At follow-up, scores for outcome measures were only used if there were <10% of items missing, and we treated outcome measures with no data as missing. We first examined descriptive statistics for each variable at baseline and used person-level means to examine between-person correlations. For time-varying measures, we also calculated within-person correlations using the *rmcorr* package [53] (Version 0.4.5) in R, version 4.2.1 (Released 2022-06-23). This package uses a modified ANCOVA framework with subjects as a between-person factor to convert the ratio of the sums of squares of within-person variability of each measure to a Pearson correlation. To examine internal consistencies, we calculated between- and within-person omega reliability [54] statistics using the omegaSEM function from the *MultilevelTools* package (Version 0.1.1).

To test the multilevel associations between selected predictor variables (e.g., mindfulness, DBT skills, and emotion dysregulation) and treatment outcomes (e.g., SUD severity and functional disability), we first disaggregated between- and within-person variability in the time-varying predictor variables [55]. Within-person variables were calculated by subtracting the person-level mean (i.e., person mean centering) from each individual data point. Between-person variables were calculated by subtracting within-person scores from grand-mean and were entered to control for between-person variability. Time and time squared were entered as continuous fixed effects measured in days and then divided by 7 to assess weeks from baseline, allowing us to examine differences in change over time and account for variability in survey completion dates between participants.

### Hypotheses 1 and 2

We ran a series of multilevel models with the *lme4* package (Version 1.1-26; [56]) for each dependent variable (SUD severity and functional disability) which included all within- and between-person variables for selected predictor variables, linear and quadratic time trends, and all demographic (e.g., age, sex, race/ethnicity) and control variables (e.g., treatment arm; baseline covariates) as fixed effects. Specifically, for Hypotheses 1a and 1b, models included within-person predictors (mindfulness, DBT skills, and emotion dysregulation), linear and quadratic time trends, and demographic and control variables (e.g., treatment arm, baseline covariates). For Hypotheses 2a and 2b, the same models additionally included between-person predictors (mindfulness, DBT skills, and emotion dysregulation). All models included a random intercept for participant and assessed whether the inclusion of the random slopes for time (linear and quadratic) and other time-varying variables improved model fit (tested one by one) using a diagonal variance-covariance structure, which was used to reduce convergence problems.

### Hypothesis 3

A similar approach was used for moderator analyses. For hypothesis 3a, time-varying moderators were disaggregated in the same way reported above to extract between-person and within-person variables (e.g., treatment acceptability, ease of use). The within-person variables were added as an interaction term with time (linear and quadratic) to assess moderation (e.g., [predictor]*time), while between-person variation, demographic variables, treatment arm, and baseline scores were added as covariates. For hypothesis 3b, certain moderators were only available as person-level (Level 2) aggregates and were not time-varying (e.g., total hours and unique days spent on iDBT). After log-transformation due to positive skew, total hours and unique days were grand-mean-centered and entered as an interaction term with time (linear and quadratic) to assess moderation (e.g., [predictor]*time). All final models included the random intercept and any random slopes that were found to improve model fit using a diagonal variance-covariance structure. Across all multilevel models, restricted maximum likelihood (REML) estimation was used to evaluate all fixed effects, but all model comparisons were evaluated using maximum likelihood estimation with the *lmerTest* (Version 3.1-3; [57]) package, which uses Satterthwaite’s degrees of freedom method. An optimizer called *optimx* (Version 2022-4.30) set to the “nlminb” method (i.e., Nonlinear Minimization subject to Box Constraints) was used in all models. All analyses were two-tailed with an alpha of .05 and reported effects are unstandardized.

### Results Preliminary Within- and Between-Person Correlations

As seen in Table 1, few significant correlations between demographic variables and other variables were found at baseline, other than a positive relationship between age and SUD severity. Many other variables demonstrated anticipated univariate associations at baseline. Between-person SUD severity was positively associated with functional disability and emotion dysregulation as well as negatively associated with mindful awareness. Between-person functional disability was also positively associated with emotion dysregulation and negatively associated with mindful awareness. These relationships were consistent at the within-person level as well. Between-person emotion dysregulation was negatively associated with mindful awareness and this extended to the within-person level as well. DBT skills were negatively associated with functional disability and emotion dysregulation at the within-person level only.

### Hypotheses 1 and 2: Within- and Between-Person Associations with iDBT Outcome

As seen in Table 2, both SUD severity and disability improved over the course of treatment. Consistent with expectations for time effects, SUD severity decreased in a non-linear fashion during iDBT (with a steeper decline closer to baseline and plateauing over time) whereas functional disability decreased in a more linear fashion. Baseline covariates for each dependent variable were strongly associated, indicating that those with higher SUD severity and disability reported more change over time. Neither demographic characteristics (i.e., age, sex, or ethnicity) nor treatment arm were associated with either outcome variable. For hypothesis 1a, within-person increases in emotion dysregulation were associated with increased SUD severity and functional disability over the course of iDBT (i.e., they impeded improvements). For hypothesis 1b, within-person mindfulness and DBT skills were not significantly associated with either outcome. For hypothesis 2a, between-person increases in mindfulness skills were associated with reduced disability over the course of iDBT. For hypothesis 2b, between-person emotion dysregulation was associated with worse outcomes, although DBT skills assessed at the within- and between-person levels were not associated with either outcome.

**Table 2.** Final models incorporating all fixed and random effects of between- and within-person skills variables predicting substance dependence and functional disability.

|  | *Model 1: Substance Dependence*<br>*(N = 72, k = 254)* |  |  |  | *Model 2: Functional Disability*<br>*(N = 72, k = 254)* |  |  |  |
| --- | --- | --- | --- | --- | --- | --- | --- | --- |
|  | *b* | *SE* | *t* | *95% CI* | *b* | *SE* | *t* | *95% CI* |
| Intercept | 7.20 | .57 | 15.58*** | 6.08, 8.32 | 15.12 | 1.23 | 13.41*** | 12.91, 17.33 |
| *Between-Person Variables* |  |  |  |  |  |  |  |  |
| Age | .03 | .02 | 1.84 | -.002, .06 | -.003 | .03 | -.11 | -.06, .06 |
| Sex (1 = female, 0 = male) | -.70 | .39 | -1.82 | -1.46, .05 | -1.16 | .78 | -1.48 | -2.70, .37 |
| Race (1 = White, 0 = other) | .05 | .36 | .15 | -.66, .77 | -.14 | .74 | -.19 | -1.59, 1.30 |
| Treatment Arm | .25 | .35 | .70 | -.44, .93 | -.13 | .67 | -.19 | -1.45, 1.19 |
| Weeks | -.40 | .08 | -4.80*** | -.57, -.24 | -.35 | .17 | -2.00* | -.69, -.007 |
| Weeks<sup>2</sup> | .02 | .01 | 3.20** | .008, .03 | .02 | .01 | 1.36 | -.008, .04 |
| Baseline DV Covariate | .83 | .06 | 14.91*** | .72, .94 | .72 | .04 | 16.35*** | .63, .81 |
| DBT Skills | .36 | .47 | .77 | -.56, 1.27 | .48 | .94 | .51 | -1.37, 2.34 |
| Emotion Dysregulation | .03 | .02 | 1.37 | -.01, .07 | .03 | .05 | .68 | -.06, .12 |
| Mindfulness Skills | -.10 | .31 | -.34 | -.71, .51 | -1.50 | .62 | -2.42* | -2.71, -.28 |
| *Within-Person Variables* |  |  |  |  |  |  |  |  |
| DBT Skills | -.14 | .69 | -.21 | -1.49, 1.21 | -2.09 | 1.29 | -1.63 | -4.61, .43 |
| Emotion Dysregulation | .08 | .02 | 3.15** | .03, .12 | .16 | .05 | 3.24** | .06, .26 |
| Mindfulness Skills | .43 | .32 | 1.34 | -.20, 1.06 | -1.24 | .67 | -1.86 | -2.55, .07 |

Note: \* $p < .05$ , \*\* $p < .01$ , \*\*\* $p < .001$ . DBT = Dialectical Behavioural Therapy. $N$ = Number of participants; $k$ = number of observations. A random intercept and all significant random slopes were included in each model using an uncorrelated diagonal setting for random effect variances.

### Hypotheses 3: Moderation of Engagement between Time and iDBT Outcome

We first assessed the time-varying cognitive engagement variables (i.e., treatment acceptability, ease of use, interface and satisfaction, and usefulness) as moderators of the relationship between time and treatment outcome, while covarying for demographics, treatment arm, and between-person average. Results of each moderator within-person interaction with time and between-person main effect are summarized in Table 3. For hypothesis 3a, only one moderation effect was found for SUD severity. There was a significant interaction between treatment acceptability and time, whereby higher within-person treatment acceptability was significantly associated with reduced SUD severity over the course of iDBT. For functional disability, there were no within-person moderation effects, but two significant between-person main effect findings emerged. Between people, higher ratings of iDBT ease of use and usefulness (indicated by lower ratings) were associated with lower functional disability score over the course treatment. For hypothesis 3b, we did not find significant interactions for either behavioral engagement variable (i.e., hours or unique days spent on iDBT), suggesting that increased time spent on iDBT did not enhance or diminish the treatment effects found for SUD severity and functional disability.

**Table 3.**
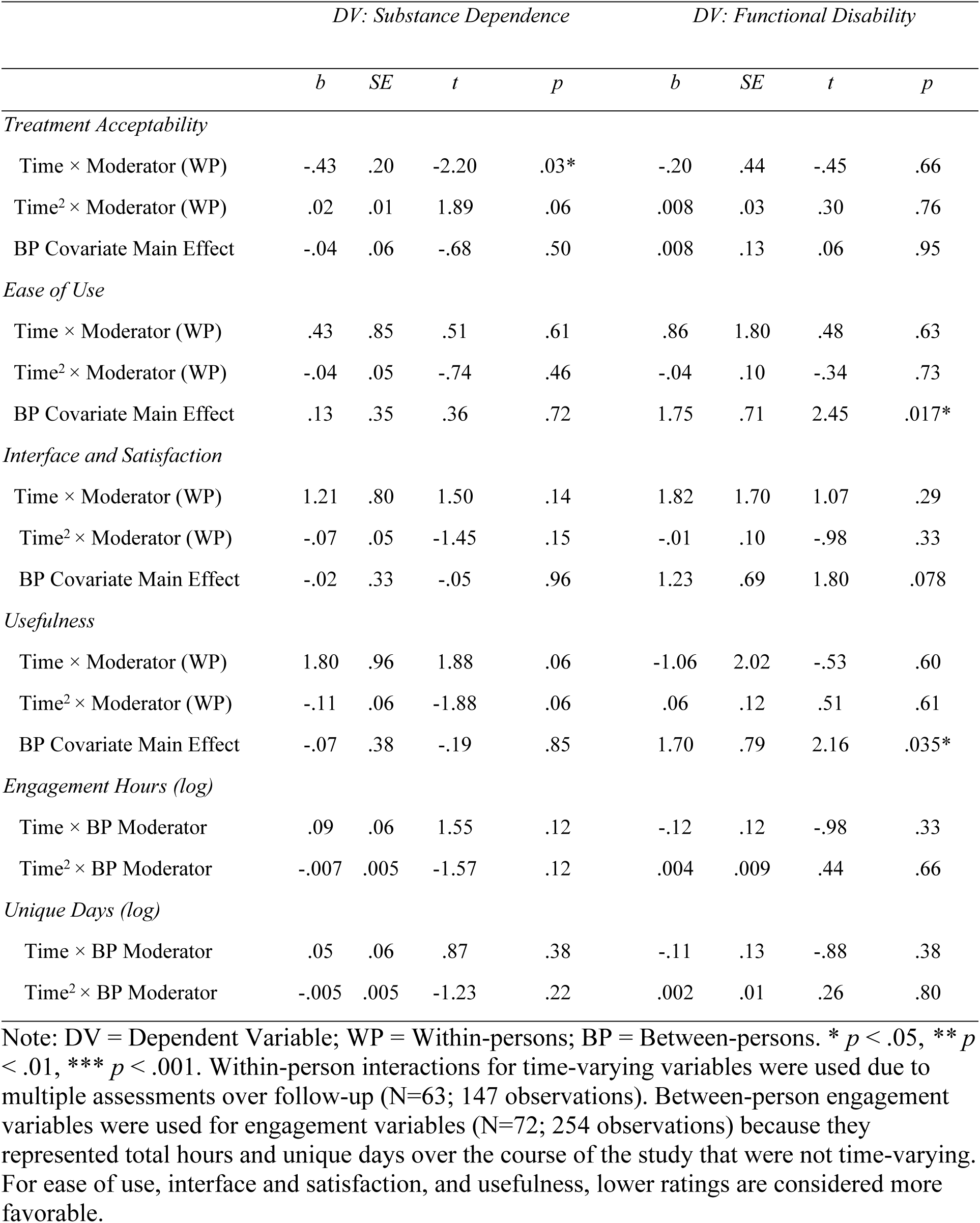
Effects of time-varying (within-person) and between-person moderation on substance dependence and functional disability as treatment outcomes.

### Sensitivity Analyses

Given the variability in findings and potential floor effects on the SUD severity and functional disability scales (i.e., several participants endorsed none to mild symptoms on each measure), we ran sensitivity analyses with depression and anxiety symptoms combined as a third outcome variable (see Table S1 in S1 Appendix). After all significant random slopes were entered and the final model was evaluated, we found similar results to the above with respect to associations with outcome. Depression and anxiety decreased non-linearly over time, with a steeper linear decline followed by plateauing (*b*_linear_ = -.58, *SE* = .20, 95% CI [-.98, -.18], *p* = .005; *b*_quadratic_ = .02, *SE* = .01, 95% CI [-.005, .05], *p* = .002). Baseline depression and anxiety was significantly associated with changes in this variable over time (*b* = .68, *SE* = .05, 95% CI [.58, .78], *p* < .001) and sex emerged as a significant covariate, indicating that male participants reported higher symptoms versus female participants (*b* = −2.98, *SE* = .89, 95% CI [-4.74, −1.23], *p* = .002).

Supporting the above main findings above, higher within-person emotion dysregulation ratings were associated with higher depression and anxiety symptoms (*b* = .21, *SE* = .06, 95% CI [.10, .32], *p* = .003) and within-person DBT skills were not associated with this outcome.

Consistent with the findings above, between-person mindfulness ratings were associated with lower depression and anxiety symptoms over the course of iDBT (*b* = −2.36, *SE* = .69, 95% [−3.73, −1.00], *p* = .001). DBT skills were not associated with depression and anxiety symptoms at the between-person level.

Similarly, we did not find effects for any within-person time-varying cognitive engagement moderators influencing the relationship between time and depression and anxiety symptoms as a treatment outcome (see Table S2 in S1 Appendix). There were, however, interaction effects between behavioural engagement and time in relation to depression and anxiety symptoms. Both higher hours spent on iDBT, as well as higher unique days, were associated with lower depression and anxiety symptoms over the course of treatment.

## Discussion

The aim of the current study was to examine potential mechanisms of treatment outcome in individuals with SUDs who received iDBT, as well as the potential moderating effect of engagement measured as both cognitive and behavioral indices. Considering the literature on both traditional and internet-delivered DBT formats, we examined whether the between- and within-person changes in DBT skills, mindfulness, and emotion dysregulation were associated with treatment response for individuals with SUDs. Moreover, we considered how time-varying indices of cognitive engagement (e.g., treatment acceptability and usability) as well as between-person behavioural engagement (e.g., hours/days spent on iDBT) enhanced or diminished these treatment effects. This study builds on prior findings by offering a more nuanced understanding of how psychological constructs fluctuate and interact during iDBT, and DBT more broadly.

Results partially supported our first and second hypotheses. Within-persons, higher emotion dysregulation (i.e., difficulties in emotion regulation) at each timepoint was associated with diminished outcomes in iDBT measured in three different ways (SUD severity, disability, and depression/anxiety). These findings support theoretical frameworks suggesting that impaired emotion regulation contributes to the development and maintenance of SUDs [6,8]. These findings are also in line with research finding improvements in emotion regulation as a predictor of change in standard DBT for a range of mental health problems, such as depression, anxiety, and self-harm [30,31]. The present study was able to extend these findings to individuals with SUDs undergoing iDBT, suggesting that changes in emotion (dys)regulation are an important variable to consider in understanding the mechanisms for favorable outcomes in this context. While we did not find within-person effects for mindfulness or DBT skills, these findings add to previous research examining these variables as potential mechanisms in standard DBT [28,29,58]. The discrepancy in findings could be due to the different format of treatment, the sample recruited, or the dose (neither of these previous studies had a focus on SUDs nor iDBT). It is possible that DBT skill acquisition requires time to integrate into daily life, and its effects may not be immediately observable within the 12-week timeframe of the current study.

Additionally, the implementation of mindfulness is difficult to monitor in asynchronous treatment and therefore is challenging to confirm use of the skills. More broadly, there is lack of research on the mechanisms of DBT treatment, making it difficult to compare findings across studies. More studies are needed to understand how people improve during DBT, especially those that consider multilevel contributions of within- and between-person variance, given the differences seen across levels in the current study.

Although within-person changes in mindfulness were not associated with treatment outcomes in iDBT for SUDs, between-person levels of mindfulness were. Individuals who had higher levels of dispositional mindfulness (assessed using [43]) also reported lower symptoms of functional disability and depression/anxiety (but not SUD severity) across treatment. Thus, mindfulness skills appear to play at least some role in the treatment outcomes of those with SUDs undergoing iDBT. This between-person effect mirrors several results found in the literature. For example, Mitchell et al. [59] found that between-person changes in mindfulness were associated with reductions in BPD symptom severity, depression, and distress in a sample of BPD individuals undergoing 20-week group DBT. In other studies, it was reported that between-person mindfulness increased over the course of standard DBT and iDBT but was not necessarily a predictor of treatment outcome [30,60,61]. Dispositional mindfulness can be seen as a protective factor because it can enhance self-awareness and reduce impulsive behaviors [62], factors that would increase behavioral control in those with SUDs. Given that we found between-rather than within-person relationships with outcomes, it suggests that more stable, trait-like individual differences in mindfulness having a stronger influence on long-term disability and depression/anxiety outcomes than momentary fluctuations [63]. Additionally, greater mindfulness abilities gained in treatment have been linked to greater gains in emotion regulation ability in at least two studies (one on mindfulness-based stress reduction [64] and another on standard DBT [58]), suggesting that it may play an important role in treatment outcome. Again, it is difficult to compare results across so few studies, emphasizing the continued need for research on mindfulness given it is a core component of DBT treatment.

With regard to moderation of treatment effects, our hypotheses were also only partially supported. While we did find that within-person increases in treatment acceptability (i.e., cognitive engagement) enhanced the relationship between time and reduction in SUD severity, there were no other within-person effects. Increased time spent on iDBT (i.e., hours/unique days) only enhanced the treatment effects for depression/anxiety symptoms, rather than SUD severity or disability. Moreover, we found that higher ease of use and usefulness ratings at the between-persons level were associated with lower functional disability (rather than within-person changes). There are some findings from the literature that align with our work. Wilks et al. [65] reported that participants who found the first module less useful and encountered technological difficulties were more likely to dropout from iDBT for SUDs, which aligns with our findings on between-person ratings of ease of use and usefulness. Another study examined the relationship between engagement on iDBT and disordered eating symptoms; however, the authors reported that neither module completion nor diary card entries were moderators of change [38]. A separate study on the same iDBT app for disordered eating found that acceptability could be cultivated through performance expectancy (i.e., perceiving that the app would be beneficial) and facilitation (i.e., app infrastructure and support; [66]). Collectively, these findings suggest that increased cognitive engagement, such as positive expectancies around trust and symptom improvement, may play a role in treatment outcome though additional iDBT trial findings would strengthen this assertion.

### Limitations and Future Directions

Despite the strengths of this study, several limitations should be considered when interpreting the findings. First, while multilevel modeling is robust to modest violations of normality, several of the variables in this study (e.g., SUD severity, functional disability, hours/unique days) were only approximately normally distributed even after transformation. This pseudo-normality may have increased residual errors, affecting the estimation of fixed effects, variance components, and interaction terms. Second, this was a secondary analysis of a single-blind RCT not originally powered to test moderation or multilevel mediation. While the analytic strategy allowed for disaggregation of within- and between-person effects and moderation by time-varying engagement variables, the sample size, particularly at the between-person level, may have been underpowered to detect small or complex interaction effects. Third, the sample consisted of treatment-seeking individuals who voluntarily enrolled in a digital intervention, which may reflect a subset of people with higher digital literacy, motivation for change, or comfort with self-guided tools. This potentially limits the generalizability of findings to broader SUD populations, particularly those with lower engagement or with more severe psychiatric comorbidities. Fourth, while we did find several significant associations with treatment outcome and some evidence of moderation, it is important to note that these relationships are still correlational. While we may hypothesize that increased skill acquisition and engagement leads to reduced SUD severity and disability, it is also plausible that reductions in symptoms facilitate greater skill learning and engagement. Therefore, causation or direction of effects cannot be assumed. Finally, the study duration of 12 weeks may have been insufficient to observe longer-term changes in skill acquisition and or symptom changes. Longer follow-up periods may be needed to assess the stability of these effects and the impact of skill acquisition and engagement on long-term treatment outcomes.

## Conclusion

This study underscores the importance of emotion regulation and mindfulness in driving treatment outcomes for individuals with SUDs undergoing iDBT. Moreover, both cognitive and behavioural engagement in the form of positive treatment perceptions and time spent on iDBT may enhance outcomes. Future research should consider the benefit of measuring these skill-building outcomes to illuminate the possible mechanisms of treatment underlying DBT in various formats as well as predictors of improved treatment response. While our findings are preliminary, there are few studies involving iDBT and therefore these results may help inform future research within different target populations and formats of DBT. Overall, iDBT for SUDs was able to enhance mindfulness and emotion regulation skills much like standard DBT and may be helpful for other diverse populations, enhancing its potential for scalability. Larger-scale investigations with longer follow up periods will meaningfully extend this foundational research.

## Data Availability

Data will be made available on request from ARD as consent for deposit in a repository was not included in our consent forms.

